# Evaluation of experiences and attitudes of patients towards patient portal enabled access to their health information or medical records – A Qualitative Study

**DOI:** 10.1101/2022.07.23.22277951

**Authors:** Preksha Machaiya Kuppanda, Judy Jenkins

## Abstract

The demand for patient centred care and patient engagement in their healthcare has driven patient portal introduction. The widespread adoption and use of patient portals, however, has been a rather slow process in the United Kingdom (UK). Hence, a limited number of studies have explored patient perceptions and experiences of general portal use which forms a foundation for successful implementation of a portal. This study, therefore, focuses on the experiences and attitudes of patients regarding use of patient portals and access to their health information. It further explores various factors perceived by patients that may influence portal use and uptake. These patient experiences were gathered through semi-structured interviews of 13 participants and the data collected was subjected to analysis using the grounded theory approach. The overall findings from this study highlights positive patient perceptions of portal use. Nevertheless, it demonstrates various areas of improvement essential to ensure successful implementation and acceptance of patient portals in the future.

**Authors summary:** Patient portals have become a globally popular tool used in the healthcare sector due to its potential to increase patient engagement which is considered essential to provide patient centred care. Similarly, the use of patient portals in the UK has increased, with different providers making this service available to patients. Patients are the key target users of patient portals, however, there is limited research that focuses on understanding patients’ perspective of using a patient portal and accessing their health information. The majority of the existing studies have either evaluated providers or healthcare professionals’ perspective of patient portal implementation or explored patient experiences of using patient portal tailored to cater individuals with specific health conditions. Therefore, our aim was to explore patients’ perception of patient portals and their experiences of accessing their health information or medical records through one. Our research has captured various factors that has influenced portal use among patients and the impact of health information access on patients and their care process. Additionally, it has identified scope for future development and discussed factors that could potentially improve patient portal implementation and drive portal use and uptake among patients.

## Introduction

There is an increased demand for patient centered care and patient engagement. This has resulted in a demand by both providers and patients to increase the role of consumers in their healthcare and decision making [1]. This and several social and human factors like healthcare expenditure, demand for home-based care, and lack of an adequate number of medical workers have led to the active implementation of patient portals [1]. Patient portals are tools that allow patients to access their health information and medical records [2-4]. These are services that are managed by a provider and are linked to a patient’s electronic health record [5, 6]. It allows patients to enter or retrieve their health information, therefore increasing patient participation [5, 7]. Patient participation in turn has the potential to improve care outcomes [5, 8].

Patient Access and myGP are patient portals used by some surgeries in the United Kingdom (UK) which provide patients with features like appointment booking, ordering repeat prescriptions, messaging, and viewing medical records [9, 10]. The NHS app is a more recent service in the country and has functions similar to the Patient Access portal while supporting additional features like the mandatory COVID-19 vaccination proof, setting organ donation preference, and checking symptoms [11, 12]. However, uptake of patient portal services, access to medical records and linked services in the UK is limited and its widespread incorporation has been a slower process compared to many first world countries [9].

Many studies associated with patient portals have included both portals and personal health records interchangeably in their study, although they vary in terms of their ownership and features [6, 13]. This has led to a failure in drawing clear differentiation between patient portals and personal health records while concluding findings [6]. Additionally, although patient portals are developed to increase patient engagement in their healthcare, there is a lack of importance given to understanding their experiences and expectations of using a portal [7]. The majority of studies have either focused on practitioners or providers perspectives of the impact of patient portal implementation [14] or have evaluated the impact of disease-specific or vendor-specific portals [9, 15]. The successful implementation of a patient portal however requires that experiences of all stakeholders, including those of patients using varying portal services be evaluated [14]. This study, therefore, aims to assess the experiences and attitudes of patients in the UK towards patient portal enabled engagement, access to medical records and linked information, and involvement in their care process.

## Methods

### Methodology

A qualitative study was conducted to evaluate the experiences of patients towards patient portal enabled engagement with their medical records and health information. A qualitative method was used as it aids in generating in-depth information of patient experiences by allowing patients to explain their perceptions in their own words rather than subjecting them to provide limited answers through a survey or structured questionnaire [16].

### Study design

#### Participants and recruitment

Adult (> 18 years old), UK Resident, and English-speaking users of either the ‘myGP’, ‘Patient Access’ or ‘NHS App’, with access to their health records or health information were targeted. Recruitment was initiated by posting an advert containing brief details of the study on various social media platforms and discussion forums. Interested participants were asked to contact the researcher by either emailing or filling an expression of interest registration form. An email containing the participant information sheet and the consent form was sent to the registered individuals who met the inclusion criteria (n=24). Individuals who consented (n=15) were further invited for a Zoom interview. 15 participants were interviewed at their convenience and 13 were included in the study (2 were excluded as they subsequently failed to meet the inclusion criteria).

#### Data collection

The information was gathered by conducting semi-structured interviews. A semi-structured interview was chosen as research states that it is the most suitable method for exploring experiences and perceptions, as it involves asking open ended questions and its flexibility allows building rapport with the participant [17]. The semi-structured interview was based on the study aim and existing literature on patient portals. The interview guide was developed based on the guidance provided in a study [18] and comprised general questions regarding the topic to aid participants to adapt to the context of the interview. It was then followed by core questions exploring the key aim of the research and supported by follow-up questions where necessary [18]. The interview was conducted via Zoom due to its ease of use, cost-effectiveness, security, data storage features, and the requirement to maintain social distancing in effect at the time of the research. [19].

#### Data analysis

The interviews were manually transcribed by the researcher, and the data were analysed using Grounded Theory (GT), as it has an inductive nature that allows for greater interpretation of healthcare experiences [20]. The constructivist GT approach was employed as it allows the researcher to engage in the creation of theories and therefore strikes a balance between participants and the researchers views in the findings [21]. Additionally, it provides the researcher with the ability to generate novel and comprehensive theories while maintaining the originality of the data collected [21, 22].

The constructivist grounded theory method comprises initial coding followed by focused coding and theoretical coding, respectively [23]. Line by line coding was employed for the initial coding of data as it provides more scope for critical evaluation of data and therefore aids in the generation of many questions to explore new concepts [24]. During the coding process, in-vivo codes comprising specific terms used by participants were used to preserve the meaning conveyed by participants. This is identified as an essential component of constructivist GT to prevent bias or extensive incorporation of the researcher’s perceptions and feelings into the participant data [24, 25].

### Ethical considerations

This study was approved by the Swansea University Medical School Research Ethics and Governance Committee (SUMS RESC project ref number 2021-0065).

## Results

### Theme 1: Patient portal and patient interaction

#### Portal service

An equal number of participants used the Patient Access and the NHS App respectively. Some had access to both the services. Participants who used Patient Access had the service for more than a year. Whereas a majority had registered with the NHS portal only in the past 6 months. The use of the NHS App was mostly driven by the need to retrieve COVID-19 vaccination proof. For example, patients stated “*The NHS app*, [used] *only recently since I heard about its introduction for the travel pass primarily*” (P10), and “*I’ve only had the NHS app since COVID*” (P07).

#### Portal features

A broad range of features were available to participants (patients) via patient portals. These features included ordering repeat prescriptions, booking appointments, and accessing medical information like health records, test reports, consultations, immunisation, and medical history. The features available were different for participants using different portal services and varied depending on the surgery. Participants highlighted this variation and one stated, “*In this* [NHS App] *also you can see consultation, but I did not see much detail* [compared to another portal service]” (P10). Another added, “*I think I’m supposed to be able to book appointments with my GP* [General Practitioner] *but I actually can’t*…*I think our GP just doesn’t use that function*” (P08).

Additionally, the commonly used features varied among participants. A participant stated, “*For patient access, the main things I used for were appointment booking at my GP*” (P07), whereas another said, “*The patient access I use it mainly for the repeat prescription and for sending requests* [medication]” (P12).

Participants displayed poor awareness of all the features available to them through the portal, some highlighted this by stating “*I can’t remember which all features it has but the main reason I use it is to order my prescription every few months due to a medical condition*” (P03), and “*I can look at repeat prescriptions but…because I haven’t had anything prescribed for a really long time, so I’m not as aware of that as a function*” (P08), respectively.

#### Patients’ portal experience

Participant perceptions of patient portals were unanimously positive. They expressed that the portal made health services and information access automated, easy, efficient, and immediate. Additionally, they described patient portals as a means to minimise elaborate conversations and unnecessary interactions with healthcare staff. One stated that “*its’s* [patient portal] *brilliant, because If I can get away with not having to ring my doctor surgery, then that is an absolute bonus for me*” (P14). Another reported, “*it* [using the portal] *was the best experience I’ve had of trying to deal with the GP and manage medicine and stuff like that*” (P08).

### Theme 2: Factors influencing patient portal use

#### General Practitioner (GP) recommendation and patient choice

In most cases, patient portals were recommended to participants by their GP for use of specific functionalities like appointment booking or medication refills. One stated “*it was then suggested* [by the surgery] *to make appointments*” (P10). Although, participants became aware of the service from recommendation by the GP surgery, the majority voiced that ultimately it was their independent choice to register to the service, by expressing “ *I was happy to like use apps all the time so I was familiar so I would have probably chosen that even though my GP hadn’t suggested it*” (P07), and “*it was my independent choice and no one forced me to use*” (P01), respectively. Additionally, participants added that the use of the service did not feel obligatory as the conventional mode of access were still available and one stated, “*it didn’t seem like I was being forced to* [use the service] *by my surgery or anything like that. So, I am still aware that there are telephone services available for people who may not have a smartphone or not want to use the app for whatever reason*” (P09). However, some participants explained that they had no choice or limited choice of services to choose from, and one stated “*my GP surgery only used the Patient Access app*…*so there was no choice, and the NHS app is the only app that gives you access to your results and the COVID pass so there was no choice. I had to get both*” (P03).

#### Perceived benefits and information need

Participants’ realisation of the potential benefits of the service aided their portal uptake and use. They recognised the ability of patient portals to enhance the speed and efficiency of their healthcare processes. They identified that portals enabled having all the services presented to them at their fingertips, thereby, making access available to them at their convenience and out of GP working hours. One mentioned, “*it’s* [portal] *a good deal, better than holding on for an hour or more trying to get through to the receptionist*” (P05).

Additionally, increased healthcare and information needs motivated use, this was evident from participants statements like, “*I was very much like wanting to know as soon as possible when the results came in, so that was sort of what spurred it* [use] *initially…I would say, I don’t use it often but if something is wrong with me at a particular time then I’ll be using it again*” (P08).

#### Pandemic and digital shift

With the COVID-19 pandemic and several healthcare services moving online, portal use, specifically the use of the NHS App among participants increased. One participant emphasised that “*It* [portal use] *is partly to do with the pandemic…was trying to get all the information because in a pandemic it makes you realise that I need to sort out and make sure that all my healthcare is okay*” (P02). Another added, “*the NHS App gives you the option to have a vaccine passport, so that’s automatically a reason to use something like that*” (P14).

### Theme 3: Patient portal enabled heath information access and its impact

#### Patient emotions

Participants had a positive experience of accessing their health records, test reports, and other health information via portal services. They were pleased to have this service available through their portal. Initial access to their health information spurred feelings of keen interest and curiosity among the participants. These emotions further developed to feeling informed, reassured, in control, less dependent, and more confident. A participant stated “*I think it is more interesting really. There was nothing, in particular, I wanted to look up*” (P05), and another added, “*I suppose it was quite freeing in a way or like it gives you a degree of independence from the doctors*” (P08). Alternatively, there were also feelings of shock and surprise among some participants after viewing old and forgotten health information and one expressed “*I really liked it, it was quite interesting and it’s quite surprising, it’s a bit strange seeing everything you’ve had wrong with you in the list and it’s quite daunting*” (P08).

#### Healthcare process

A majority of the participants expressed that portal enabled access to health information and records had a positive impact on their healthcare process, while some stated that it made little or no significant difference to their care. One explained that “*It’s not that just because you can see your health information, disease or whatever condition, it doesn’t mean that you then become aware of your health…I would say it hasn’t made any difference by having access*” (P10). Many participants highlighted that access to their health information made them newly aware of their medical history or allowed them to recall forgotten health information, with one mentioning “*long time ago I had an allergy to penicillin, that was recorded which I myself had forgotte*n” (P13). Additionally, access to health records allowed patients to easily compare and identify previous treatments that have worked. It further helped by bridging any communication gaps and language barriers and enabled patients to be more proactive, involve in shared decision making, and make informed healthcare choices. One participant stated “*Some of the things the doctors said I didn’t fully understand. But with the app, I can look at it myself*” (P12), a second added “*I find that really reassuring I’ve got the level same access level as they* [doctors] *do*” (P14).

#### Conventional versus portal supported health information access

While expressing how portal access made a difference from the conventional method of receiving and accessing health information, participants emphasised that portal access is comparatively faster, and serves immediate information needs. They highlighted that it makes information access easier and meaningful with all health information available at one place, thus, preventing scattering of health records and aiding the generation of longitudinal health data. Additionally, participants appreciated the ability of the service to allow them to access and interact with their health data at a time and place of their convenience. These views are evident from participants stating, “*It’s much more efficient, it’s much easier, it’s quicker, it’s done in my time and in my speed at my convenience*” (P05), “*It’s more convenient rather than getting updates from different places like mail messages or whatever*” (P01), and “*Through the app, you can just access it on your own terms, no one is trying to prompt you*” (P02), respectively.

#### Patient perceived drawbacks

Participants identified varying threats of having access to health information and records via their patient portal. Key concerns included an obsession of viewing records, and the potential risk of self-diagnosis. One pointed that “*I think having access to your own records will lead to people jumping to wrong conclusions about their records, whereas on the other hand, it might require them to see a GP but because you have access to your records you might be less inclined*” (P02).

Additionally, participants acknowledged data security and privacy issues. A significant number, however, had little or no privacy concerns. This was due to their confidence in either the service provided by the NHS, their devices security system, or both. Many believed that a strong password was key to ensuring data security. This was evident with participants stating, “*I have no concerns because…I think National Health is being quiet, the data protection and all that, they take that, you know seriously (sic)*.” (P13), and “*it’s just a case of being able to make a good password*” (P09). On the other hand, a few were apprehensive of potential hacking but were willing to make trade-offs, either due to the absence of confidential information present within their records or due to their perceived benefits of patient portals. A participant expressed “*I am not worried because I don’t think there is anything on there that I am worried about anyone seeing or using it*” (P02).

### Theme 4: Patient portal adaptability and ease of use

#### Information interpretation

Most of the participants reported that the health information presented to them via their portal was comprehensible and provided detailed information of their diagnosis or treatment. Some believed that the interpretability of the information depended on the type of report, and level of individuals medical knowledge. This was further reflected among participants as they expressed that although they managed to understand the information presented, they experienced some level of difficulty in interpreting the medical terms and stated that they relied on colloquial and supported explanations to interpret the data. For example, a participant stated, “*I obviously don’t understand the medical term but if the word normal is used that I will think that is okay*” (P04). Many also mentioned relying on google and other resources to research and understand complex information.

#### Technological literacy

None of the participants expressed having trouble in navigating through the given patient portal, however, they voiced their concern regarding the limited accessibility of portals for the disabled and the older population with poor technological literacy. One participant stated, “*To me, I would say there are no downsides, but I can see that as a problem for people who have no help, no knowledge of how to use the system*” (P10), and another added, “*I think this* [patient portal] *is more towards the younger generation than the older ones*” (P13).

### Theme 5: Expectations and future developments

#### Patient expectation

Several participants had no initial expectations from patient portals and accessing health records, as they registered to the service out of curiosity or to use it specifically for repeat prescriptions or appointment booking. A participant expressed “*I had no expectation, to begin with, I didn’t know what to expect so I think it has met my expectation”* (P13), and another stated, “*At first, I was curious, and I’ve accessed some fairly old documents going back to 1993… It was an interesting read, and I am glad I saw those letters*” (P05). They were fascinated to see various features, their health information, and medical history in their patient portal. All participants considered the service reliable in general, but this perception varied for different features. Additionally, one pointed that “*it is still dependent on human input, so it is as reliable as you consider a human*” (P02), and another added that the service reliability depended on the GP surgery providing it. Furthermore, some had suggestions for future improvement. A participant emphasised this by stating “*So far it has met my expectations. Obviously going forward there is a lot of room for improvement*” (P12).

#### Future developments

Some of the key suggestions of portal improvements among participants included obtaining more information access via patient portals, and incorporation of additional features that can further aid their healthcare process. Many voiced the need for establishing consistency of service and portal features offered across GP surgeries and increased promotion of portal services to enhance portal use and adoption. These were explicitly stated by participants as “*I don’t understand why all GPs don’t use it. I think if all surgeries used it, it would be a lot easier for people to understand because it would just be one process for everyone*” (P07), and “*I did speak to a few of my friends and not many people seem to be using it* [patient portal], *I don’t think it is heavily promoted. So, I think marketing a bit more*” (P13).

## Discussion

The participants in this study used the NHS App, Patient Access service, or both. None of them used the myGP service: this could be due to poor promotion of the service in the participants practice as highlighted in a study by Ryan et al [26] or due to promotion of alternative services. A broad range of features were available to participants through the NHS and the Patient Access portals, including ordering repeat prescriptions, booking appointments, viewing medical records, test results, and accessing consultation documents. However, the features available and used by each participant varied, depending on the portal service used and their registered General Practitioner (GP) surgery. The variation of features within the same portal service is due to the surgery being responsible for deciding what portal services are accessible to their patients [27, 28]. Additionally, most of the participants displayed poor awareness of various features within their patient portal, therefore, leading to non-use. This results when there is a lack of communication and guidance provided to patients by the providers [29, 30]. Patients are usually provided with details of setting up an account and logging in, whereas information given on features is not elaborate [4]. Portal introduction must, therefore, be accompanied by adequate patient training and guidance to enable effective use of the service [4].

Majority of the participants in this study revealed that their GP recommended the respective portal service, which is known to play a significant role in patient portal adoption and use [7]. Although portal services were recommended, participants expressed having an open choice to register for the service. Hence, their continued portal use was influenced by their voluntary interest and perceived benefits of the service, which are considered as key factors essential for long term use of a portal [31-33]. Additionally, the use of the NHS App among participants was strongly influenced by its feature which allowed them to access their COVID-19 vaccination proof. This outcome is in accordance with a study [29], which highlights that in many cases patient portal use is a result of a reactive process to either a policy or a process.

Overall, among the participants, booking appointments and ordering repeat prescriptions were the most widely used features within a portal, whereas, accessing health records and test results were a result of curiosity or response to the availability of the service. This is consistent with previous research findings [9, 34]. Alternatively, studies by Rodriguez [35] and Moll et al. [36] reported that accessing medical records and viewing test results were the most used features of the portal. This difference in findings could be a result of varying medical and health information needs among portal users. For example, in this study the appreciation of the ability to access health records and information via a portal was directly proportional to the healthcare needs of the participants. This finding is consistent with previous studies which have noted that individuals’ health, discharge, and medication status influence portal use [29,36-40].

Irrespective of their health status, participants had positive experiences of viewing their health information, medical records, or test results. They reported feeling confident, in control, aware, and informed of their health and care process. This response is consistent with previous studies [34, 36, 41]. Furthermore, participants expressed feeling reassured by being able to access information which earlier only their healthcare practitioners had access to. Some also cited that portal access helped bridge language and communication barriers with the providers. This equivalent access and enhanced information communication prevent patients from feeling powerless and ignored [42], which in turn can aid in safeguarding their emotional well-being which can otherwise be negatively influenced by caring neglect [43].

In addition to expressing these emotions, participants noted several benefits of accessing their health information and records. They expressed becoming aware of their medical history, and allergies which they previously had no knowledge of or had forgotten. This awareness aids in ensuring patient safety by minimising the risk of patient allergies and history being ignored during treatments [38]. Access to medical history further allowed participants to compare different treatments, identify progress and patterns, and make informed choices in their ongoing or upcoming care. A similar benefit was voiced in a study by Fisher, Bhavnani, and Winfield [40].

Furthermore, accessing health information and records via a portal was deemed easy, quick, and efficient by the participants. They valued the ability of portals to make seeking emergency medical assistance easier, with all necessary health information and history easily accessible. Individuals travelling frequently considered this as one of the most beneficial aspects of electronic access, since it allowed them to easily share health information with different care providers and healthcare services. This in turn can enhance care coordination, save time and resources otherwise spent on conducting repetitive consultations and tests [44]. Additionally, participants reported that electronic access to their health information helped to keep them on track of their healthcare timeline by keeping them up to date regarding their treatments, immunisation, medication, and therefore ensured receiving medical attention when necessary. This outcome is useful as in most cases there is a lack of clarity regarding who is responsible for test result dissemination, thereby resulting in serious safety implications for patients due to the potential failure to follow up [45].

Participants in this study identified patient portals as more beneficial compared to the conventional modes of accessing health information as they not only catered for their health information needs but also provided multiple services. This could be one of the drivers to portal use as a study [39] suggests that access to records alone is not viewed as a useful service by patients but this feature supported by options like appointment booking, messaging, and ordering prescription intrigued patients. Additionally, it allowed having all resources and data in one place and prevented scattering of information. Participants further emphasised their appreciation regarding the possibility of accessing information at a time and pace suitable to them. Similar admiration is reflected among patients in a study by Zanaboni et al [46]. Furthermore, participants were not very concerned regarding their ability to interpret the health data presented to them. Although many reported difficulties in understanding medical terminologies which is considered essential for interpreting medical information [47], the ability to view their health information in accordance with their convenience aided their data interpretation. Some reported looking up the internet and researching as methods used to aid their interpretation. On the other hand, some responses reflected that they did not make an effort to understand the information in depth, instead referred to informal and simple terms to ensure the reports are normal.

The majority of the participants displayed awareness regarding data privacy, but it was one of the least expressed concerns. Among users of the NHS app, this response was majorly due to trust in services provided by the NHS, therefore, suggesting the influence of product branding which was demonstrated as one of the strengths of the NHS app by Beaney et al [48]. The lack of concern was also influenced by participants perception of the level of sensitivity of their medical information within their portal. On the other hand, some participants were willing to make trade-offs. This aligns with findings of a study [49] which noted that irrespective of their privacy concerns, patients are keen to use patient portals and access their health information as they believe that the benefits of the service outweigh any potential harm or drawbacks.

No participants in this study exhibited personal negative experiences of portal use. Nevertheless, they expressed their concerns about how a portal could negatively impact a certain set of people. The most expressed concern was, patient portals potential to stem disparity by limiting access to old, disabled, and individuals with poor health and technological literacy. This risk holds true in a study [31] that reported lower portal acceptance among the older and vulnerable groups as a result of poor health literacy or resistance to change. Alternatively, another study [50] identified that older adults registered to one or more portals irrespective of their technological literacy, therefore, highlighting their interest in using a portal. This suggests that increased provider encouragement, support, and training can prevent these individuals from being alienated from the service [32, 51]. Furthermore, there were concerns that access to health information and medical history via patient portals might result in paranoia and obsession among some individuals. Similarly, physicians in a study [52] expressed concern that access to health information could fuel hypochondria among users.

Despite these general concerns, the services offered by these portals met majority of the participants expectations. The access to health information feature was beyond the initial expectation of most of the participants. Nevertheless, many expressed keen interest in obtaining more health information and details of their medical records. This demand is persistent among patients across various studies [53-55], therefore, promising future success of portal implementation and adoption.

## Limitations

This study has some limitations. Firstly, this research explores the perception of patient portal users only in the UK. This therefore could limit generalisability of these findings to patient portal users on a global scale, as patient perceptions of portal use are strongly influenced by service provider and product branding, which vary across geographical boundaries. However, findings suggesting approaches to drive portal use, uptake, and health information access by patients are generic and independent of the service provider and can, thus be applied globally. Secondly, the process used to recruit participants through social media adverts could have limited participation of individuals inactive on these platforms or those with minimal access to the internet. Thirdly, some Zoom interviews were interrupted by the individuals’ video conferencing set up which could have restricted them from providing elaborate answers. The impact of these on the study results however are minimal, as 13 participants which is considered as the minimum number required to achieve saturation in an interview-based study were included [56]. Additionally, Zoom offered the advantage of effectively accessing participants while managing the limited time and resources that were available for the study.

## Conclusion

Findings from this study contribute towards understanding patients’ experiences of accessing health information through patient portals. It was evident from the findings that portal use among patients is influenced by the service provider and portal features. Whereas their perceptions of accessing health information are influenced by their health situation and information needs. Although these perceptions were collectively positive, there is scope for future development. A key area of improvement is the need for establishing consistency of portal service offered across surgeries. This can aid familiarity and usability of the service, therefore, avoiding confusion among patients. Additionally, there is a need for increasing awareness of the service, its available features and providing patients with the necessary support in the form of training and encouragement to enable uniform access and use.

## Data Availability

The data associated with this study included original participant interview recordings and transcripts. As per the ethical approval from Swansea University Medical School Research Ethics and Governance Committee, the interview recordings were deleted post publication of my Master's results. The interview transcripts retention is approved for a further period of 2 years (up to May 2024), therefore, these transcripts cannot be made publicly available. For more information on data availability please contact me at

## References

1. Zarcadoolas C, Vaughon WL, Czaja SJ, Levy J, Rockoff ML. Consumers’ perceptions of patient-accessible electronic medical records. Journal of medical Internet research. 2013;15(8):e168.

2. Nøst TH, Faxvaag A, Steinsbekk A. Participants’ views and experiences from setting up a shared patient portal for primary and specialist health services-a qualitative study. BMC Health Services Research. 2021 Dec;21(1):1–9.

3. Hoogenbosch B, Postma J, Janneke M, Tiemessen NA, van Delden JJ, van Os-Medendorp H. Use and the users of a patient portal: cross-sectional study. Journal of medical Internet research. 2018;20(9):e262.

4. Lee JL, Williams CE, Baird S, Matthias MS, Weiner M. Too many don’ts and not enough do’s? A survey of hospitals about their portal instructions for patients. Journal of general internal medicine. 2020 Apr;35(4):1029–34.

5. Irizarry T, Dabbs AD, Curran CR. Patient portals and patient engagement: a state of the science review. Journal of medical Internet research. 2015 Jun;17(6):e148.

6. Kruse CS, Bolton K, Freriks G. The effect of patient portals on quality outcomes and its implications to meaningful use: a systematic review. Journal of medical Internet research. 2015;17(2):e44.

7. Ryan BL, Brown JB, Terry A, Cejic S, Stewart M, Thind A. Implementing and using a patient portal: a qualitative exploration of patient and provider perspectives on engaging patients. Journal of innovation in health informatics. 2016 Jul 4;23(2):534–40.

8. Laurance J, Henderson S, Howitt PJ, Matar M, Al Kuwari H, Edgman-Levitan S, et al. Patient engagement: four case studies that highlight the potential for improved health outcomes and reduced costs. Health Affairs. 2014 Sep;33(9):1627–34.

9. Mohammed MA, Montague J, Faisal M, Lamming L. The value of a patient access portal in primary care: a cross-sectional survey of 62,486 registered users in the UK. Universal Access in the Information Society. 2020 Nov;19(4):855–72.

10. myGP. Book doctor’s appointment [Internet]. London: iPlato Healthcare Ltd; 2020 [cited 2021 Aug 5]. Available from: https://www.mygp.com/user/

11. Best J. The NHS App: opening the NHS’s new digital “front door” to the private sector. BMJ. 2019 Oct 29;367.

12. Nelson SJ, Allkins S. Technology in healthcare: the NHS app. British Journal of Cardiac Nursing. 2020 Jan 2;15(1):1–3.

13. Zhao JY, Song B, Anand E, Schwartz D, Panesar M, Jackson GP, et al. Barriers, facilitators, and solutions to optimal patient portal and personal health record use: a systematic review of the literature. AMIA annual symposium proceedings; 2017.American Medical Informatics Association; 2017. p. 1913–22

14. Kooij L, Groen WG, van Harten WH. Barriers and Facilitators Affecting Patient Portal Implementation from an Organizational Perspective: Qualitative Study. Journal of medical Internet research. 2018;20(5):e183.

15. Sääskilahti M, Ahonen R, Timonen J. Pharmacy Customers’ Experiences of Use, Usability, and Satisfaction of a Nationwide Patient Portal: Survey Study. Journal of medical Internet research. 2021 Jul 16;23(7):e25368

16. LaVela SL, Gallan AS. Evaluation and measurement of patient experience. Patient Experience Journal. 2014;1(1):28–36.

17. Al-Busaidi ZQ. Qualitative research and its uses in health care. Sultan Qaboos University Medical Journal. 2008;8(1):11–19.

18. DeJonckheere M, Vaughn LM. Semistructured interviewing in primary care research: a balance of relationship and rigour. Family Medicine and Community Health. 2019;7(2):e000057

19. Archibald MM, Ambagtsheer RC, Casey MG, Lawless M. Using zoom videoconferencing for qualitative data collection: perceptions and experiences of researchers and participants. International Journal of Qualitative Methods. 2019;18: 1–8

20. Sbaraini A, Carter SM, Evans RW, Blinkhorn A. How to do a grounded theory study: a worked example of a study of dental practices. BMC medical research methodology. 2011;11:128.

21. Alemu G, Stevens B, Ross P, Chandler J. The use of a constructivist grounded theory method to explore the role of socially-constructed metadata (Web 2.0) approaches. Qualitative and Quantitative Methods in Libraries. 2017;4(3):517–40.

22. Chong CH, Yeo KJ. An overview of grounded theory design in educational research. Asian Social Science. 2015;11(12):258–68.

23. Tie YC, Birks M, Francis K. Grounded theory research: A design framework for novice researchers. SAGE open medicine. 2019;7:1–8

24. Charmaz K. Constructing grounded theory: A practical guide through qualitative analysis. SAGE; 2006. Chapter 3, Coding in grounded theory practice; p. 42–71

25. Chiovitti RF, Piran N. Rigour and grounded theory research. Journal of advanced nursing. 2003;44(4):427–35.

26. Ryan M, Marlow L, Forster A, Ruwende J, Waller J. Offering an app to book cervical screening appointments: A service evaluation. Journal of medical screening. 2020;27(2):85–9.

27. Patient Access. View your medical record [Internet]. Leeds: EMIS; n.d [cited 2021 Nov 27]. Available from: https://support.patientaccess.com/medical-record-viewer/can-i-see-my-medical-record-in-patient-access

28. NHS. GP surgery appointments [Internet]. [cited 2021 Nov 27]. Available from: https://www.nhs.uk/nhs-app/nhs-app-help-and-support/appointments-and-online-consultations-in-the-nhs-app/gp-surgery-appointments/

29. Powell KR, Myers CR. Electronic patient portals: patient and provider perceptions. On-Line Journal of Nursing Informatics. 2018;22(1).

30. Walker, J., Leveille, S., Bell, S., Chimowitz, H., Dong, Z., Elmore, J. G., et al. OpenNotes after 7 years: patient experiences with ongoing access to their clinicians’ outpatient visit notes. Journal of medical Internet research. 2019 22(4): e13876.

31. Wildenbos GA, Peute L, Jaspers M. Facilitators and barriers of electronic health record patient portal adoption by older adults: a literature study. Stud Health Technol Inform. 2017 ;235(235):308–12.

32. Mishuris RG, Stewart M, Fix GM, Marcello T, McInnes DK, Hogan TP, et al. Barriers to patient portal access among veterans receiving home-based primary care: a qualitative study. Health Expectations. 2015;18(6):2296–305.

33. Elers P, Nelson F. Improving healthcare through digital connection? Findings from a qualitative study about patient portals in New Zealand. Australian Journal of Primary Health. 2018;24(5):404–8.

34. Alpert JM, Morris BB, Thomson MD, Matin K, Brown RF. Identifying how patient portals impact communication in oncology. Health communication. 2019;34(12):1395–403.

35. Rodriguez ES. Using patient portals to increase engagement in patients with cancer. Semin Oncol Nurs. 2018;34(2):177–83.

36. Moll J, Rexhepi H, Cajander Å, Grünloh C, Huvila I, Hägglund M, et al. Patients’ experiences of accessing their electronic health records: national patient survey in Sweden. Journal of medical Internet research. 2018;20(11):e278.

37. Ancker JS, Osorio SN, Cheriff A, Cole CL, Silver M, Kaushal R. Patient activation and use of an electronic patient portal. Informatics for Health and Social Care. 2015;40(3):254–66.

38. Ordaz OH, Croff RL, Robinson LD, Shea SA, Bowles NP. Optimization of primary care among Black Americans using patient portals: qualitative study. Journal of medical Internet research. 2021;23(6):e27820.

39. Hannan A. Providing patients online access to their primary care computerised medical records: a case study of sharing and caring. Journal of Innovation in Health Informatics. 2010;18(1):41–9.

40. Fisher B, Bhavnani V, Winfield M. How patients use access to their full health records: a qualitative study of patients in general practice. Journal of the Royal Society of Medicine. 2009;102(12):539–44.

41. Mold F, de Lusignan S, Sheikh A, Majeed A, Wyatt JC, Quinn T, et al. Patients’ online access to their electronic health records and linked online services: a systematic review in primary care. British Journal of General Practice. 2015;65(632):e141–51.

42. Rathert C, Mittler JN, Banerjee S, McDaniel J. Patient-centered communication in the era of electronic health records: What does the evidence say?. Patient education and counseling. 2017;100(1):50–64.

43. Reader TW, Gillespie A. Patient neglect in healthcare institutions: a systematic review and conceptual model. BMC health services research. 2013 Dec;13(1):1–5.

44. Baudendistel I, Winkler EC, Kamradt M, Brophy S, Längst G, Eckrich F, et al. Cross-sectoral cancer care: views from patients and health care professionals regarding a personal electronic health record. European journal of cancer care. 2017;26(2):e12429.

45. Thomas J, Dahm MR, Li J, Georgiou A. Can patients contribute to enhancing the safety and effectiveness of test-result follow-up? Qualitative outcomes from a health consumer workshop. Health Expectations. 2021;24(2):222–33..

46. Zanaboni P, Kummervold PE, Sørensen T, Johansen MA. Patient use and experience with online access to electronic health records in Norway: results from an online survey. Journal of medical Internet research. 2020;22(2):e16144.

47. Freise L, Neves AL, Flott K, Harrison P, Kelly J, Darzi A, et al. Assessment of patients’ ability to review electronic health record information to identify potential errors: cross-sectional web-based survey. JMIR formative research. 2021;5(2):e19074.

48. Beaney P, Odulaja A, Hadley A, Prince C, Obe RC. GP online: turning expectations into reality with the new NHS app. British Journal of General Practice. 2019;69(681):172–3.

49. Vodicka E, Mejilla R, Leveille SG, Ralston JD, Darer JD, Delbanco T, et al. Online access to doctors’ notes: patient concerns about privacy. Journal of medical Internet research. 2013;15(9):e2670.

50. Lockwood MB, Dunn-Lopez K, Pauls H, Burke L, Shah SD, Saunders MA. If you build it, they may not come: modifiable barriers to patient portal use among pre-and post-kidney transplant patients. JAMIA open. 2018;1(2):255–64.

51. Lyles, C. R., Sarkar, U., Ralston, J. D., Adler, N., Schillinger, D., Moffet, H. H., … & Karter, A. J. (2013). Patient–provider communication and trust in relation to use of an online patient portal among diabetes patients: the diabetes and aging study. Journal of the American Medical Informatics Association, 20(6), 1128–1131.

52. Witry MJ, Doucette WR, Daly JM, Levy BT, Chrischilles EA. Family physician perceptions of personal health records. Perspectives in health information management/AHIMA, American Health Information Management Association. 2010;7(Winter).

53. Dendere R, Slade C, Burton-Jones A, Sullivan C, Staib A, Janda M. Patient portals facilitating engagement with inpatient electronic medical records: a systematic review. Journal of medical Internet research. 2019;21(4):e12779.

54. Van den Bulck SA, Hermens R, Slegers K, Vandenberghe B, Goderis G, Vankrunkelsven P. Designing a patient portal for patient-centered care: cross-sectional survey. Journal of medical Internet research. 2018;20(10):e9497.

55. Sulieman L, Steitz B, Rosenbloom ST. Analysis of employee patient portal use and electronic health record access at an academic medical center. Applied clinical informatics. 2020 May;11(03):433–41.

56. Vasileiou K, Barnett J, Thorpe S, Young T. Characterising and justifying sample size sufficiency in interview-based studies: systematic analysis of qualitative health research over a 15-year period. BMC medical research methodology. 2018;18(1)

